# Adverse Events Following SARS-CoV-2 mRNA Vaccination Among Patients with Inflammatory Bowel Disease

**DOI:** 10.1101/2021.03.30.21254607

**Authors:** Gregory J. Botwin, Dalin Li, Jane Figueiredo, Susan Cheng, Jonathan Braun, Dermot P.B. McGovern, Gil Y. Melmed

## Abstract

Patients with immune-mediated inflammatory diseases (IMID) such as inflammatory bowel disease (IBD) on immunosuppressive and biologic therapies were largely excluded from SARS-CoV-2 mRNA vaccine trials. We thus evaluated post-mRNA vaccination adverse events (AE) in 246 vaccinated adults with IBD participating in a longitudinal vaccine registry. In general, AE frequency was similar to that reported in the general population. As in the general population, AE were more common among younger patients, and those with prior COVID-19. We additionally found that AE were less common in individuals receiving biologic therapy. Those with IBD and other IMID on these commonly prescribed therapies can be reassured that the AE risk is likely not increased, and may be reduced, while on biologics.

## Introduction

Patients with immune-mediated inflammatory diseases (IMID) such as inflammatory bowel disease (IBD) receiving immunosuppressive and biologic therapies were underrepresented in SARS-CoV-2 mRNA vaccine trials.^1, 2^ Thus, the safety and efficacy of these vaccines in this population is largely unknown. Concerns about adverse events (AE), or reactogenicity, were reported in 70% of those with IBD hesitant to receive SARS-CoV-2 vaccination.^3^ We aimed to clarify the AE frequency and severity in this vulnerable vaccine-eligible population.

## Methods

We enrolled adults with IBD planning to receive, or who already received SARS-CoV-2 vaccination into the Coronavirus Risk Associations and Longitudinal Evaluation-IBD (Corale-IBD) study, a prospective, nationwide registry. We longitudinally surveyed post-vaccination symptoms after each vaccine dose among those who received at least 1 dose of a 2-dose series of either BNT162b2 (Pfizer-BioNTech) or mRNA-1273 (Moderna) vaccines using a web-based survey administered 8 days following each dose, or at the time of enrollment in the registry if it exceeded 8 days after the last vaccine dose. Symptoms were assessed across multiple organ systems, and were graded by severity (i.e. mild, moderate, or severe impact on activities of daily living, or requiring hospitalization as defined in Baden et al^2^) and duration (<2 days, between 2 and up to 7 days, or more than 7 days). We analyzed post-vaccination symptoms by age, sex, disease classification, vaccine type, and medication classes using students T-test (R version 3.5.1). Corticosteroids, immunomodulators, biologics, and Janus kinase (JAK) inhibitors were classified as “immune-modifying” therapies. Anti-tumor necrosis factor (TNF)-α, anti-integrin (vedolizumab), anti-interleukin (IL)-12/23, and small-molecule JAK inhibitors were classified as “biologic” therapies. Multivariable regression was used to examine the association of predictors and AE risk while adjusting for potential confounders. The study protocol was approved by the Cedars-Sinai institutional review board. All study participants provided informed consent.

## Results

As of March 22, 2021, 246 subjects (mean age 47.4y; 57% female) had completed an AE assessment following at least 1 mRNA vaccine dose (**Table**). One hundred forty-one (57%) received BNT162b2 (Pfizer), and 105 (42.7%) received mRNA-1273 (Moderna). IBD diagnosis included Crohn’s disease (67%) and ulcerative/indeterminate colitis (33%). Nine subjects (3.7%) reported prior COVID infection. At the time of vaccination, 50 (20.3%) were not receiving any immune-modifying therapies. The proportions of patients receiving systemic corticosteroids, thiopurine monotherapy, anti-tumor necrosis factor-α (TNF) monotherapy, anti-TNF with concomitant thiopurine, vedolizumab, ustekinumab, and tofacitinib were 6.9%, 2.4%, 28.0%, 8.5%, 13.4%, 16.7%, and 1.6%, respectively.

**Table:** Participant characteristics

|  | Crohn's<br>disease<br>(N=165) | Ulcerative/indetermin<br>ate colitis<br>(N=81) | Total<br>(N=246) |
| --- | --- | --- | --- |
| **Age at Enrollment** |  |  |  |
| Mean (SD) | 48.1 (16.1) | 46.1 (14.4) | 47.4 (15.5) |
| Median [Min, Max] | 45.0 [20.0,<br>85.0] | 43.0 [22.0, 79.0] | 44.0 [20.0,<br>85.0] |
| **Sex** |  |  |  |
| Male | 56 (33.9%) | 33 (40.7%) | 89 (36.2%) |
| Female | 96 (58.2%) | 43 (53.1%) | 139 (56.5%) |
| Prefer not to answer | 1 (0.6%) | 0 (0%) | 1 (0.4%) |
| **Self Reported Race** |  |  |  |
| White | 139 (84.2%) | 62 (76.5%) | 201 (81.7%) |
| Black or African American | 5 (3.0%) | 0 (0%) | 5 (2.0%) |
| Asian | 2 (1.2%) | 6 (7.4%) | 8 (3.3%) |
| Native Hawaiian or other Pacific<br>Islander | 0 (0%) | 0 (0%) | 0 (0%) |
| American Indian/ Alaska Native | 0 (0%) | 1 (1.2%) | 1 (0.4%) |
| Multiple | 3 (1.8%) | 1 (1.2%) | 4 (1.6%) |
| Other | 3 (1.8%) | 6 (7.4%) | 9 (3.7%) |
| Prefer not to answer | 1 (0.6%) | 0 (0%) | 1 (0.4%) |
| **Self-Reported Hispanic Ethnicity** |  |  |  |
| Hispanic/LatinX | 6 (3.6%) | 1 (1.2%) | 7 (2.8%) |
| Non-Hispanic/LatinX | 147 (89.1%) | 75 (92.6%) | 222 (90.2%) |
| Prefer not to answer | 0 (0%) | 0 (0%) | 0 (0%) |
| **BMI** |  |  |  |
| Mean (SD) | 25.5 (6.39) | 25.6 (5.74) | 25.6 (6.17) |
| Median [Min, Max] | 23.7 [15.5,<br>67.3] | 23.6 [18.1, 44.5] | 23.7 [15.5,<br>67.3] |
| **Ever Smoked** |  |  |  |
| Yes | 21 (12.7%) | 16 (19.8%) | 37 (15.0%) |
| No | 100 (60.6%) | 51 (63.0%) | 151 (61.4%) |
| **Currently Smokes** |  |  |  |
| Yes | 0 (0%) | 0 (0%) | 0 (0%) |
| No | 21 (12.7%) | 16 (19.8%) | 37 (15.0%) |
| **Month of Enrollment** |  |  |  |
| Dec, 2020 | 3 (1.8%) | 1 (1.2%) | 4 (1.6%) |
| Jan, 2021 | 19 (11.5%) | 8 (9.9%) | 27 (11.0%) |
| Feb, 2021 | 52 (31.5%) | 33 (40.7%) | 85 (34.6%) |
| Mar, 2021 | 91 (55.2%) | 39 (48.1%) | 130 (52.8%) |
| **Vaccine Type** |  |  |  |
| BNT162 (Pfizer/BioNtech) | 93 (56.4%) | 48 (59.3%) | 141 (57.3%) |
| mRNA-1273 (Moderna/NIH) | 72 (43.6%) | 33 (40.7%) | 105 (42.7%) |
| **Prior COVID Infection** |  |  |  |
| No | 146 (88.5%) | 70 (86.4%) | 216 (87.8%) |
| Yes | 4 (2.4%) | 5 (6.2%) | 9 (3.7%) |
| **Age at IBD Diagnosis** |  |  |  |
| Mean (SD) | 28.3 (15.4) | 30.7 (12.9) | 29.1 (14.6) |
| Median [Min, Max] | 25.0 [4.00,<br>85.0] | 28.0 [8.00, 64.0] | 26.0 [4.00,<br>85.0] |
| **Duration (years) of IBD** |  |  |  |
| Mean (SD) | 19.5 (14.6) | 15.6 (11.9) | 18.2 (13.8) |
| Median [Min, Max] | 16.0 [0, 61.0] | 13.0 [0, 49.0] | 15.0 [0, 61.0] |
| **Prior Vaccine Adverse Reaction** |  |  |  |
| Yes | 7 (4.2%) | 4 (4.9%) | 11 (4.5%) |
| No | 157 (95.2%) | 74 (91.4%) | 231 (93.9%) |
| Not sure | 1 (0.6%) | 3 (3.7%) | 4 (1.6%) |
| **Sulfasalazine/mesalamine** |  |  |  |
| Yes | 25 (15.2%) | 33 (40.7%) | 58 (23.6%) |
| No | 132 (80.0%) | 46 (56.8%) | 178 (72.4%) |
| **Budesonide** |  |  |  |
| Yes | 4 (2.4%) | 2 (2.5%) | 6 (2.4%) |
| No | 153 (92.7%) | 77 (95.1%) | 230 (93.5%) |
| **Oral/parenteral steroids** |  |  |  |
| Yes | 12 (7.3%) | 5 (6.2%) | 17 (6.9%) |
| No | 145 (87.9%) | 74 (91.4%) | 219 (89.0%) |
| **6MP/azathioprine monotherapy** |  |  |  |
| Yes | 4 (2.4%) | 2 (2.5%) | 6 (2.4%) |
| No | 153 (92.7%) | 77 (95.1%) | 230 (93.5%) |
| **Methotrexate monotherapy** |  |  |  |
| Yes | 0 (0%) | 0 (0%) | 0 (0%) |
| No | 157 (95.2%) | 79 (97.5%) | 236 (95.9%) |
| **Anti-TNF without 6MP/AZA/MTX** |  |  |  |
| Yes | 51 (30.9%) | 18 (22.2%) | 69 (28.0%) |
| No | 106 (64.2%) | 61 (75.3%) | 167 (67.9%) |
| **Anti-TNF + 6MP/AZA/MTX** |  |  |  |
| Yes | 12 (7.3%) | 9 (11.1%) | 21 (8.5%) |
| No | 145 (87.9%) | 70 (86.4%) | 215 (87.4%) |
| **Anti-integrin** |  |  |  |
| Yes | 25 (15.2%) | 8 (9.9%) | 33 (13.4%) |
| No | 132 (80.0%) | 71 (87.7%) | 203 (82.5%) |
| **IL12/23 inhibitor** |  |  |  |
| Yes | 37 (22.4%) | 4 (4.9%) | 41 (16.7%) |
| No | 120 (72.7%) | 75 (92.6%) | 195 (79.3%) |
| **JAK inhibitor** |  |  |  |
| Yes | 1 (0.6%) | 3 (3.7%) | 4 (1.6%) |
| No | 156 (94.5%) | 76 (93.8%) | 232 (94.3%) |
| **5-ASA, oral or rectal** |  |  |  |
| Yes | 25 (15.2%) | 33 (40.7%) | 58 (23.6%) |
| No | 132 (80.0%) | 46 (56.8%) | 178 (72.4%) |
| **Other** |  |  |  |
| Yes | 8 (4.8%) | 3 (3.7%) | 11 (4.5%) |
| No | 149 (90.3%) | 76 (93.8%) | 225 (91.5%) |
| **Any steroids** |  |  |  |
| No | 137 (83.0%) | 69 (85.2%) | 206 (83.7%) |
| Yes | 20 (12.1%) | 10 (12.3%) | 30 (12.2%) |
| **Any Biologic** |  |  |  |
| No | 31 (18.8%) | 37 (45.7%) | 68 (27.6%) |
| Yes | 126 (76.4%) | 42 (51.9%) | 168 (68.3%) |
| **Any immunomodulator** |  |  |  |
| No | 134 (81.2%) | 67 (82.7%) | 201 (81.7%) |
| Yes | 23 (13.9%) | 12 (14.8%) | 35 (14.2%) |
| **No immune-modifying therapies** |  |  |  |
| No | 136 (82.4%) | 50 (61.7%) | 186 (75.6%) |
| Yes | 21 (12.7%) | 29 (35.8%) | 50 (20.3%) |

The overall AE frequency was 39% after dose 1 (D1), and 62% after dose 2 (D2) (**Figure**). Localized injection-site reactions were reported in 38% after D1, and in 56% after D2. The most common systemic AE included fatigue/malaise (reported by 23% after D1, and 45% after D2), headache/dizziness (14% after D1, 34% after D2) and fever/chills (5% after D1, 29% after D2). Gastrointestinal symptoms were reported by 6% after D1, and 11.5% after D2. These frequencies are similar to the clinical trial reports as well as data from a distinct health-care worker study conducted at the same site and using similar AE capture methods.^1, 4^

**Figure:**
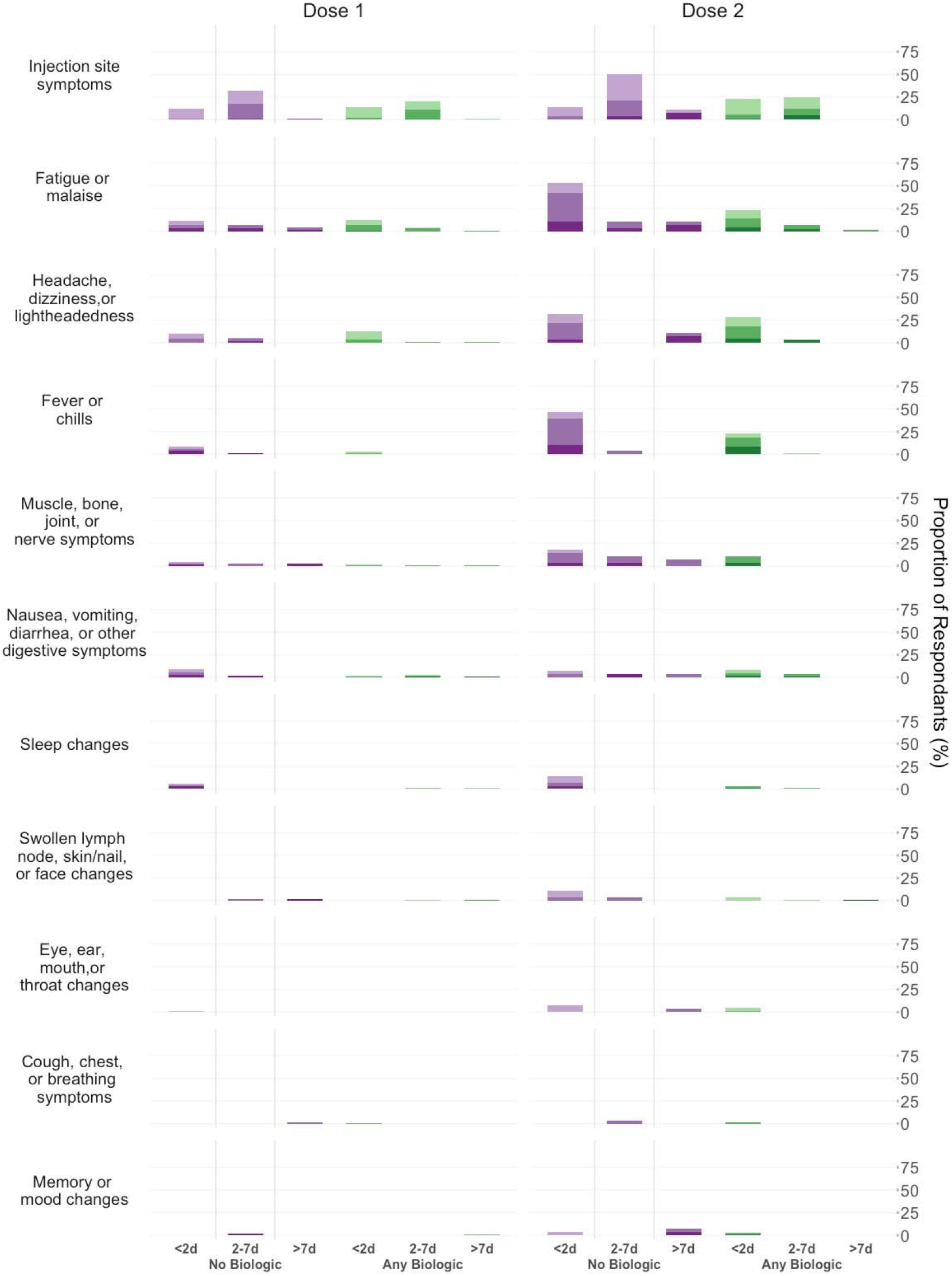
Adverse events after each vaccine dose stratified by biologic use (purple: no biologic; green: any biologic). AEs are graded by severity (darker colors represent more severe AE). AE are also tracked by duration. (“Biologic” refers to use of anti-TNFα, anti-integrin, anti-IL12/23, or JAK-inhibitor)

The vast majority of AE were non-severe. The most common severe symptom after D1 was fatigue/malaise (3%); other severe symptoms were reported by 2% or fewer subjects. The most common severe symptoms after D2 included fatigue/malaise (10%), fever/chills (8%), and headache (8%). Most symptoms resolved in less than 2 days except for injection site reactions, which mostly resolved within 7 days. Twenty-seven severe AE (defined as preventing daily sactivity) were reported by 16 subjects after D1, and 53 severe AE were reported by 24 subjects after D2. Three subjects were hospitalized after D1: two for fatigue/malaise, and one with gastrointestinal symptoms. Two subjects were hospitalized after D2: one for gastrointestinal symptoms and fever, and one for headache, fever, and fatigue.

AE frequency was higher among subjects younger than 50 years relative to older subjects (47% vs 29% after D1, p=0.011; 73% vs 45% after D2, p=0.003), among those with a prior COVID history (78% vs 37% after D1, p=0.04), and among those with ulcerative colitis relative to those with Crohn’s disease (78% vs 55% after D2, p=0.038); no significant differences by disease type were observed after D1. Subjects receiving biologic therapy were less likely to report AE than those not on biologic therapies (36% vs 47% after D1, p=0.17; 54% vs 82% after D2, p=0.013). Similarly, subjects receiving any immune-modifying therapy were less likely to report AE than those not on any immune-modifying therapies (37% vs 48% after D1; p=0.22; 54% vs 86% after D2; p=0.012). No significant differences in any AE frequency were seen based on sex, vaccine type, or history of prior infusion reaction.

We assessed the association of biologic therapies on AE after adjusting for age, disease type, and prior COVID status. Only age was associated with AE after D1 (OR 0.97, 95% CI 0.96–0.99; p=0.015), suggesting reduced AE risk with each year of advancing age. Significant AE associations after D2 included age (OR 0.97, 95% CI 0.94–0.99; p=0.018) and biologic status (OR 0.32, 95% CI 0.10-0.94; p=0.049), suggesting a reduced AE risk among biologic recipients, independent of age. Neither disease type nor prior COVID were retained in the adjusted models, although only 9 patients had previous COVID.

## Discussion

Concerns for AE are an important contributor to persistent vaccine hesitancy, particularly among those with chronic immune-mediated diseases.^3^ We found that AE incidence in patients with IBD was generally similar to previously reported rates^1, 2, 4, 5^ Furthermore, severe AE or those lasting more than 7 days were very uncommon.

Consistent with the results of post-vaccine responses in general populations, we identified younger age and prior COVID-19 infection as independent predictors of AE to mRNA vaccines among patients with IBD.^1, 2, 4, 5^ Notably, individuals on any biologic therapy were less likely to experience AE, particularly after D2. Those with IBD and other IMID on these commonly prescribed therapies can be reassured that the AE risk is likely not increased, and may actually be reduced, while on biologics. It remains to be seen whether biologics are associated with blunted post-vaccination serologic responses, and whether post-vaccine serologic responses correlates with AE frequency or severity. A single mRNA vaccine dose in organ transplant recipients receiving anti-metabolite maintenance immunosuppression therapy were less likely to develop anti-spike serologic responses 2 weeks after vaccination.^6^ However, organ transplant recipients have historically had lower vaccine response rates compared to those with IMID.^7^

Our study is limited by lack of racial/ethnic diversity, and potential recall bias. However, our results have direct and immediate clinical relevance. Our findings that biologic therapies may attenuate vaccine-related AE can be considered reassuring to patients taking these therapies.

## Data Availability

Data are available in the manuscript and supporting files.

## Acknowledgements

We thank all who have contributed to the CORALE-IBD study:

Keren Appel, Andrea Banty, Edward Feldman, Christina Ha, Rashmi Kumar, Susie Lee, Shervin Rabizadeh, Theodore Stein, Gaurav Syal, Stephan Targan, Eric Vasiliauskas, David Ziring

Brigid Boland, Aline Charabaty, Michael Chiorean, Erica Cohen, Rebecca Fausel, Ann Flynn, David Fudman, Arash Horizon, Jason Hou, Caroline Hwang, Mark Lazarev, Donald Lum, Mark Mattar, Mark Metwally, Arthur Ostrov, Nimisha Parekh, Laura Raffals, Swapna Reddy, Emilie Regner, Sarah Sheibani, Corey Siegel, John Valentine, Doug Wolf, Ziad Younes

Jennifer Davis, Phillip Debbas, Mary Hanna, Elizabeth Khanishian, Melissa Hampton, Justina Ibrahim, Emebet Mengesha, Angela Mujukian, Ashley Porter, Valeriya Pozdnyakova, Aura Ruiz, Shane White, Cindy Zamudio

James Beekley, Sarah Contreas, Joseph Ebinger, Ergueen Herrera, Amy Hoang, Sandy Joung, Nathalie Nguyen, Sarah Sternbach, Nancy Sun, Min Wu

